# A systematic review and meta-analysis on effectiveness and safety of traditional Chinese medicine combined with EGFR-TKIs in treating advanced non-small cell lung cancer

**DOI:** 10.1101/2022.06.11.22276127

**Authors:** Shanyang Su, Yanling Huang, Wenjia Li, Xiang Lu, Xiaowan Chen, Jihong Zhou

## Abstract

**Introduction:** EGFR-TKIs have been shown in multiple clinical trials to be effective in first-line therapy, but acquired resistance is inevitable regardless of the recent effect. EGFR-TKIs combined with TCM comprehensive therapy have shown unique effects in clinical randomized trials to increase the efficacy of radiotherapy and chemotherapy and gene-targeted drugs, reduce their adverse reactions, reduce the chance of recurrence and metastasis, enhance the immune recognition and killing of lung cancer cells, and reverse the multi-drug resistance of lung cancer cells. Therefore, our study aims to clarify the clinical efficacy, safety and benefit of EGFR-TKIs combined with TCM in the treatment of advanced non-small cell lung cancer.

**Methods and Analysis:** We will search four international electronic databases (PubMed, Cochrane Library, EMBASE and Web of Science) and four Chinese electronic databases (CNKI, VIP, WanFang, China Biology Medicine) to retrieve relevant literature. We only included studies from launch until publication in May 2022. The primary outcomes will include the Response Evaluation Criteria in Solid Tumours、 progression-free survival and effectiveness rate。 Secondary outcomes will include Karnofsky functional status scale, 1-year survival rate, and grading scale for common toxicities of anticancer drugs(WHO). Two reviewers will conduct independent research selection, data extraction, data synthesis and quality assessment. The assessment of bias risk and data synthesis will be conducted using Review Manager 5.3 software. Improved Jadad scale will be used to evaluate the quality of evidence.

**Ethics and Dissemination:** Because this systematic review will be conducted based on published research, there is no ethical approval requirement. The findings of this systematic review will be published in a peerreviewed journal.

## 1. Introduction

Primary bronchial lung cancer, referred to as lung cancer, is a malignant tumor that originates from the bronchial mucosa or glands. Lung cancer has insidious onset, asymptomatic in the early stage, and clinical manifestations such as cough, sputum blood or hemoptysis, weight loss, and hoarseness in the middle and late stages. Therefore, most patients are found to be in advanced stages, there is no chance of early surgical cure, survival and quality of life are also significantly reduced, and some studies have shown that the five-year survival rate of lung cancer patients is only 19.8%.^[1]^ The latest cancer data published by WHO’s International Agency for Research on Cancer in 2021 shows that lung cancer remains the cancer with the highest mortality rate.^[2]^ The social burden of lung cancer is increasing.^[3]^ With the successful development of molecularly targeted drugs, the therapeutic level of NSCLC has been continuously improved. One of the current NSCLC targets of greatest concern is the epidermal growth factor receptor (EGFR).^.[4]^ Epidermal growth factor-tyrosine kinase inhibitors (EGFR-TKIs) are widely used in the clinical. treatment of mid- and advanced NSCLCs and are the first-line treatment for mutation-positive non-small cell lung cancer.

Enhancing the efficacy of EGFR-TKIs and delaying their resistance is one of the current research priorities of targeted therapies. Targeted therapy has broken the long-term situation of poor chemotherapy efficacy and obvious toxicity. Although EGFR has been shown in several clinical trials to be effective in first-line therapy, almost all patients will eventually develop acquired resistance to EGFR-TKIs, regardless of recent efficacy.^[5,6]^ Chemotherapy combined with EGFR-TKIs has been shown to prolong PFS by approximately five months compared with EGFR-TKIs alone.^[7, 8]^ However, the combination of first-line EGFR-TKIs with chemoradiotherapy will limit the options for second-line treatment after tumor progression. EGFR-TKIs alone can be used as a first-line treatment, and after tumor progression, chemoradiation and chemotherapy can also be used as an effective second-line regimen. Mutations in exon T790M are mostly related to drug-resistant mutations, with 50-70% of patients having mutations in the EGFR gene T790M. Osimitinib is the third generation of EGFR-TKIs, and osimitinib has shown significant efficacy in the treatment of NSCLCs that are resistant to erlotinib, gefitinib, or alfati due to the development of T790M mutations.^[9,10]^ However, the current mechanism of oxitinib resistance and the choice of treatment after drug resistance are still unclear, and when the third generation of EGFR-TKIs is resistant, there is no new generation of EGFR-TKIs to choose from. Secondary drug resistance puts treatment in a bind.

In addition to combination chemotherapy and timely replacement with a new generation of EGFR-TKIs,^[11, 12]^ Chinese decoctions enhance autoimmune function in patients, improve their quality of life, and reduce the occurrence of adverse reactions.^[13-15]^ EGFR-TKIs combined with TCM therapy have shown unique effects in clinical randomized trials to increase the efficacy of radiotherapy and chemotherapy and gene-targeted drugs, reduce their adverse reactions, reduce the chance of recurrence and metastasis, enhance the immune recognition and killing of lung cancer cells, and reverse the multi-drug resistance of lung cancer cells.^[16]^ Longer PFS have been reported in studies with EGFR-TKIs combined with first-line treatment of TCM for NSCLC, particularly for exon 21 mutations (L858R).^[17]^ There are currently no studies on the efficacy and safety of EGFR-TKIs combined with TCM in the treatment of advanced non-small cell lung cancer. Our study aims to determine the clinical efficacy, safety and benefits of EGFR-TKI in combination with TCM for advanced non-small cell lung cancer, or whether they may be harmful to patients. It is a therapeutic strategy for advanced non-small cell lung cancer in primary clinical diagnosis and treatment, especially for delaying acquired resistance to EGFR-TKI.

## 2. Materials and methods

### 2.1 Study registration

The study protocol has been registered in PROSPERO (registration number: CRD42022332404). Evaluation reports will be conducted in accordance with the preferred reporting items in the systematic review and meta-analysis guidelines.

### 2.2 Study design

#### 2.2.1 Type of participants

Have a definite pathological or cytological diagnosis of NSCLC and the stage is IIIB-IV. Genetic testing shows positive for mutations in the EGFR driver gene.

#### 2.2.2 Type of interventions

ALL randomised controlled trials of NSCLC in combination with Traditional Chinese medicine plus EGFR-TKIs will be included. Patients in the treatment group will be given EGFR-TKIs supplemented with traditional Chinese medicine therapeutic measures, which are administered orally only, whereas patients in the control group will be given EGFR-TKIs treatment only. The dosages and courses are not limited in our studies.

#### 2.2.3 Types of outcome measures

The primary outcomes will include the Response Evaluation Criteria in Solid Tumours、 progression-free survival and effectiveness rate。 Secondary outcomes will include Karnofsky functional status scale, 1-year survival rate, and grading scale for common toxicities of anticancer drugs(WHO).

#### 2.2.4 Inclusion and exclusion criteri

Studies that met all of the following requirements will be included:

① Have a definite pathological or cytological diagnosis of NSCLC and the stage is IIIB-IV;
② The research type is a randomized controlled clinical trial;
③ The age, gender, race, etc. of the research subjects are not limited;
④ The language of the literature is Chinese and English;
⑤ The experimental group was EGFR-TKIs (Gefitinib, or Erlotinib, or Icotinib, or Afatinib, or Dacomitinib, or Osimertinib) plus middle Medical treatment measures (limited to oral administration); the control group is Gefitinib, or Erlotinib, or Icotinib, or Afatinib, or Dacomitinib, or Osimertinib.

Studies that met one of the following requirements will be excluded:

① The research data is incomplete or erroneous, or the complete data cannot be obtained;
② Repeated publication and repeated detection of documents;
③ The sample size of the experimental group was less than 30 cases;
④ The Chinese medicine of the intervention is non-oral;
⑤ Case reports, animal experiments, qualitative studies, reviews or review articles.

### 2.3 Literature search strategy

We will comprehensively search four international electronic databases (PubMed, Cochrane Library, EMBASE and Web of Science) and four Chinese electronic databases (CNKI, VIP, WanFang, China Biology Medicine) to retrieve relevant literature. To avoid duplication of ongoing clinical trials, we will search the following 2 trial registries for identifying relevant studies: the China Clinical National Trial Registry www.chictr.org.cn/index.aspx) The Institute of Health’s In-Progress Trial Registry (www.clinicaltrials.gov), the World Health Organization International Clinical Trials Registry Platform (www.who.int/trialsearch). We only included studies from launch until publication in May 2022. These studies must be published in English or Chinese. The literature search will be structured around search terms for non-small cell lung, search terms for EGFR-TKIs (Gefitinib, Erlotinib, Icotinib, Afatinib, Dacomitinib, Osimertinib, Gefitinib, Erlotinib, Icotinib, Afatinib, Dacomitinib, Osimertinib),search terms for traditional Chinese medicine, search terms for randomized controlled trials, and adjusted for each database as needed. References to included studies will also be screened for further inclusion of material. PubMed’s detailed search strategy is shown in Table 1.

**Table 1.**
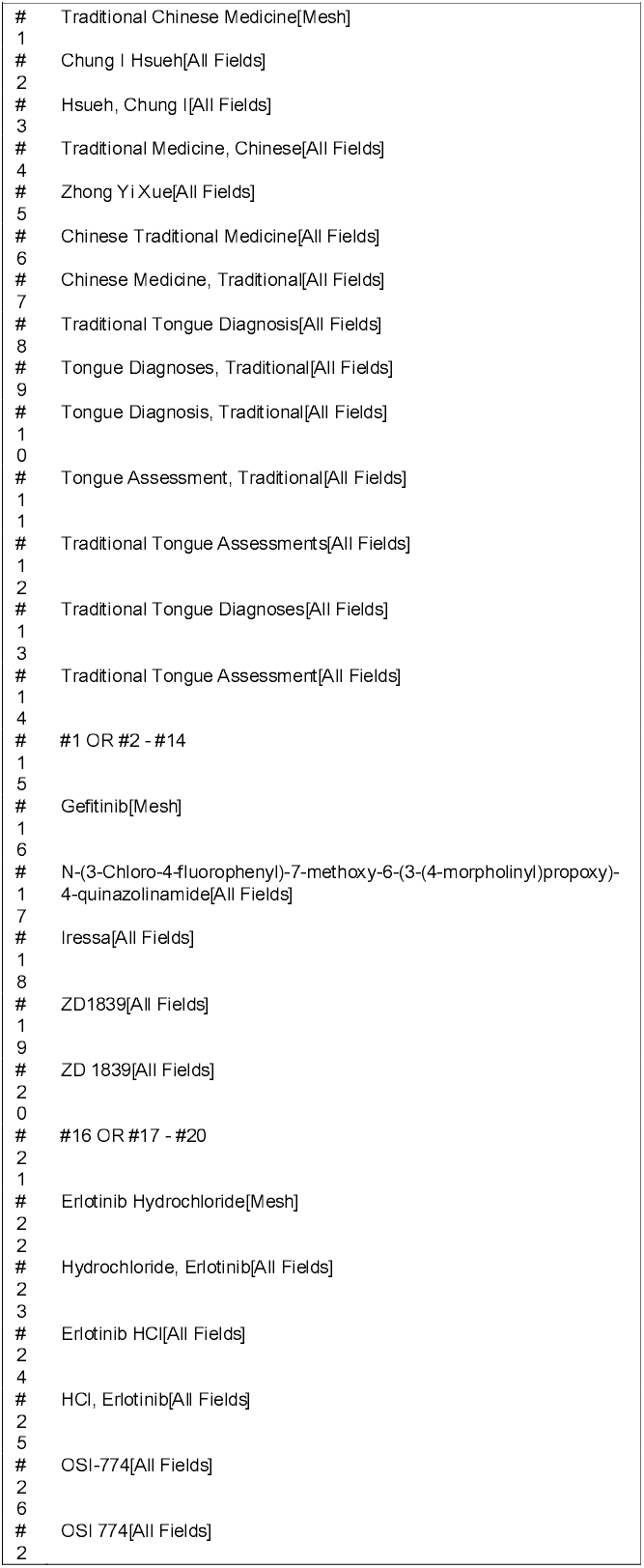
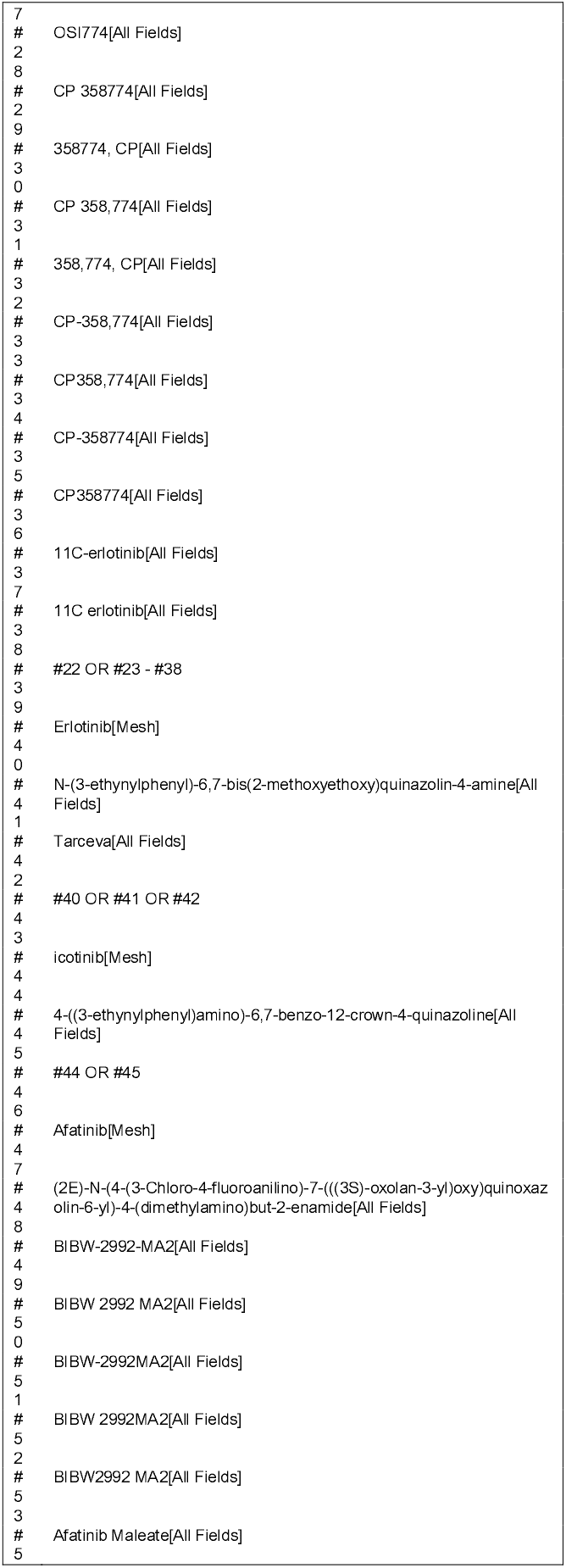
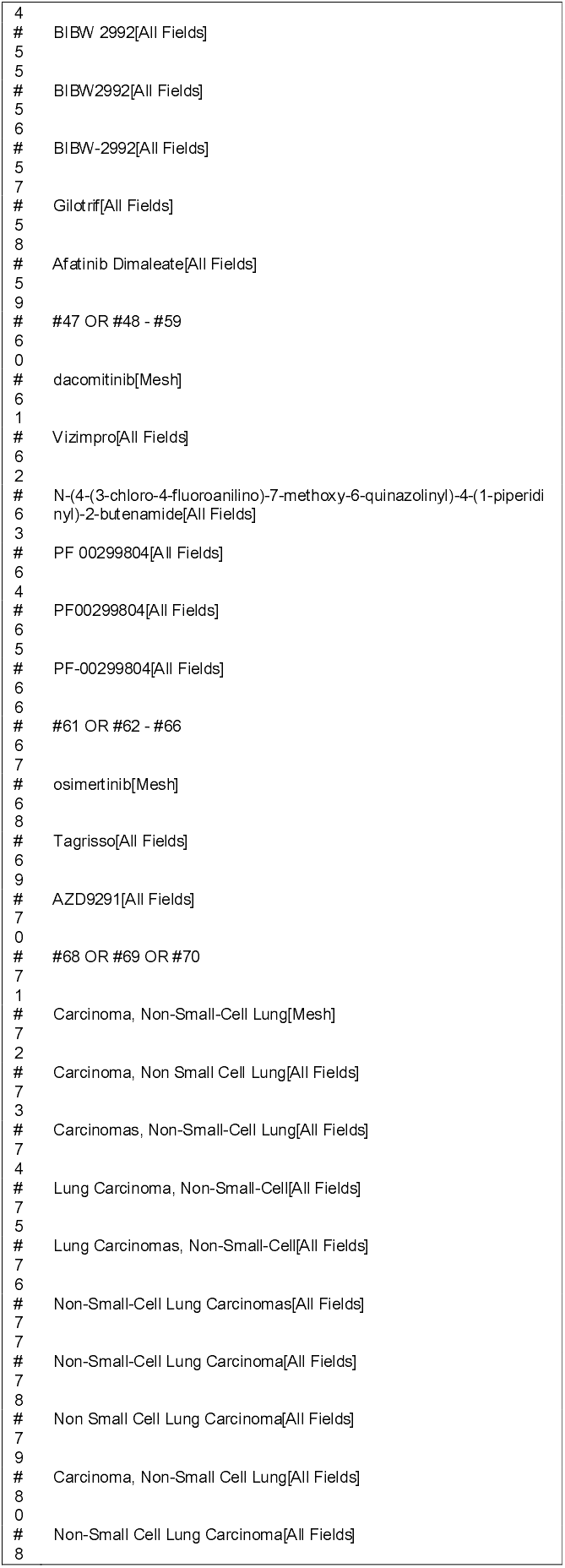
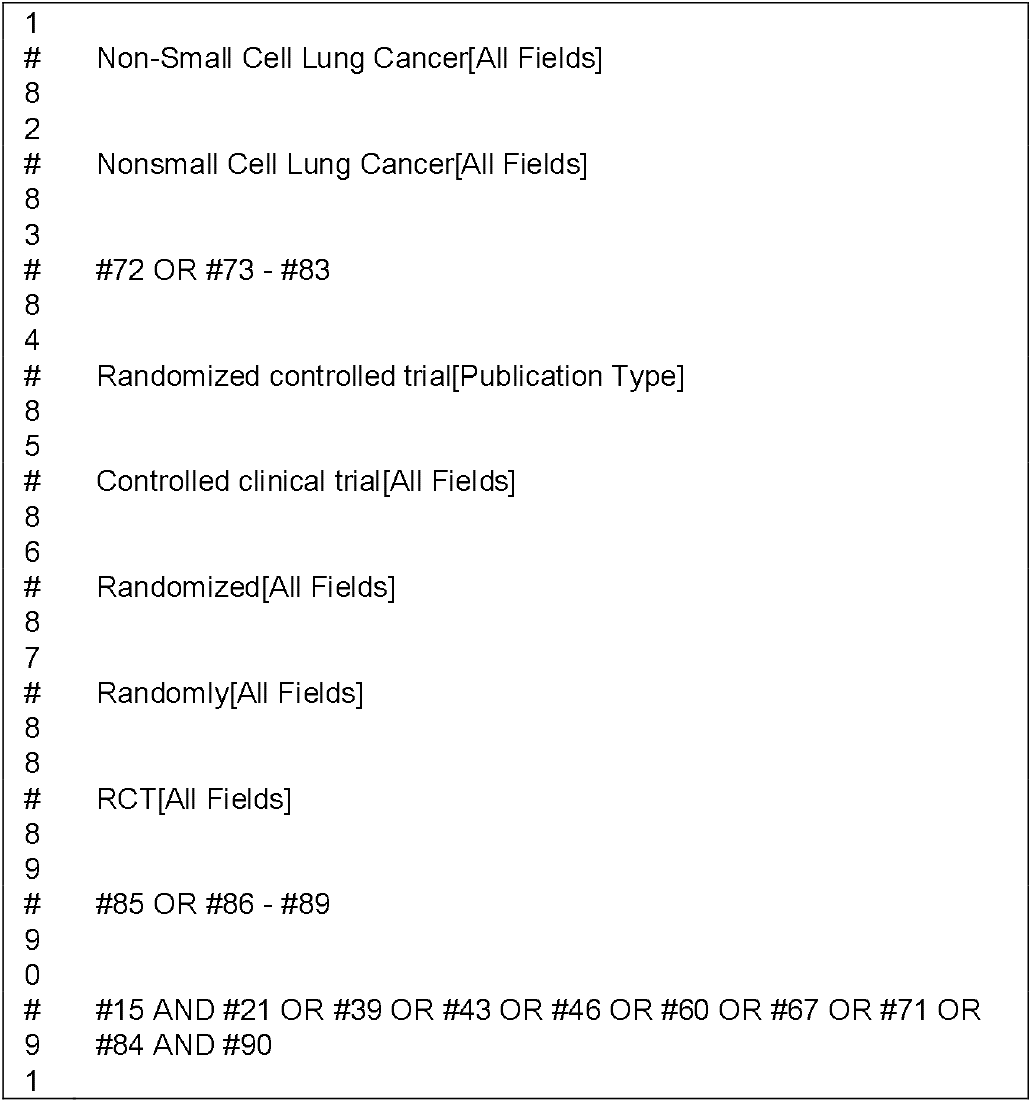
Search strategy in PubMed.

### 2.4 Study selection

In the first step of the data processing process, the retrieval strategy retrieves the titles and abstracts of all studies that are filtered for relevance, and the titles and abstracts of all studies that are clearly irrelevant will be discarded. If the results are not obviously irrelevant, the full text will be downloaded. In the second step, two members of the evaluation panel (Shanyang Su and Yanling Huang) will independently assess the eligibility of the study using the redefined inclusion and exclusion criteria. In addition, for studies that meet the inclusion criteria, reviewers will read the entire article to ensure that the entire study meets the criteria and are prepared to extract relevant information. Any disagreements on whether to include a particular study will be resolved through discussions between reviewers. (with a third author if necessary) to identify and resolve. For data on the missing part, we will contact the study authors for the missing data. The flowchart of all study selection procedures is shown in Figure 1

**Figure 1:**
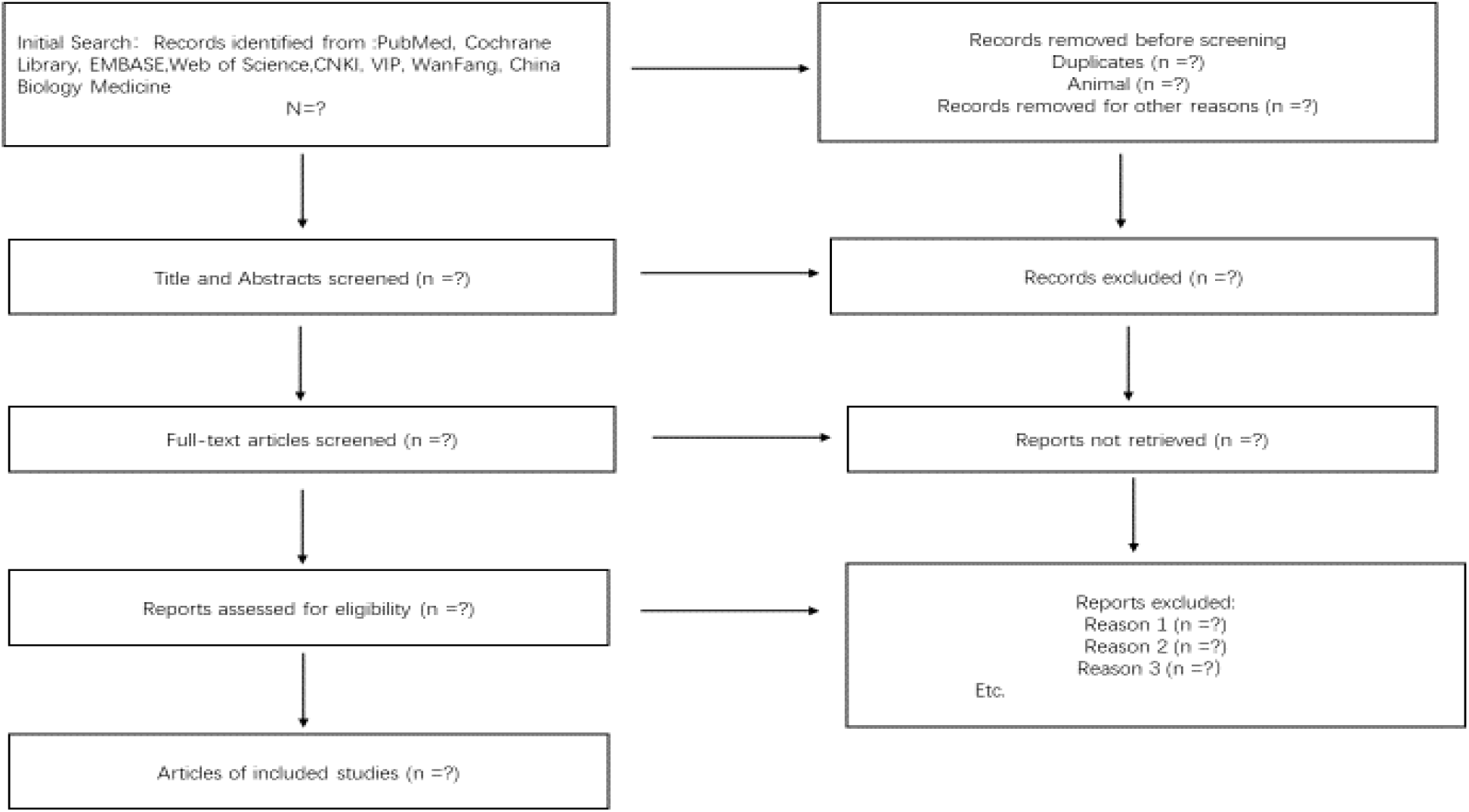
Flowchart of study selection procedures.

### 2.5 Data extraction

Two authors(Shanyang Su and Yanling Huang) will independently extract data. Any disagreement will be resolved by discussion until consensus is reached or by consulting a third author. The following data will be extracted: author, year of publication, country where the study was conducted, study period, total number of people included in the study, names of EGFR-TKIs, combination of Chinese medicines, Response Evaluation Criteria in Solid Tumours、 progression-free survival and effectiveness rate 、 Karnofsky functional status scale, 1-year survival rate, and grading scale for common toxicities of anticancer drugs(WHO). If important data are missing, we will ask the study authors to provide the number of missing data.

### 2.6 Risk of bias Assessment

There will be 2 reviewers involved in the quality assessment process, and any major disagreements will be resolved by discussion to define the final set of included studies. Two reviewers will independently assess the risk of bias in included studies by considering the following characteristics: randomization sequence generation, treatment allocation concealment, blinding method, completeness of outcome data, selective outcome reporting, and other sources of bias. Besides, the Cochrane Collaboration’s risk of bias assessment tool will be used to assess the individual included studies’ quality.

### 2.7 Data synthesis

We will use Review Manager 5.3 software to carry out the quantitative synthesis if the included studies are sufficiently homogeneous. Mean difference or standardized means difference will be used for continuous data. Risk ratio (RR) will be used for the analysis of dichotomous data. Both we will give a 95% confidence interval (CI). The heterogeneity test will be tested with χ^2^ when P>0. 1, I^2^ in 0 ∼ 50%, we will use a fixed-effect model for meta-analysis, when P≤0. 1, I^2^ in 50% to 75%, will be used random effects model for meta-analysis. Descriptive analysis will be performed when I^2^ is between 75% and 100%.

#### 2.7.1 Analysis of subgroups

A subgroup analysis of some of the plans will be conducted: efficacy of different EGFR-TKIs combine with Chinese Medicine (e.g., first-generation EGFR-TKIs, second-generation EGFR-TKIs, third-generation EGFR-TKIs) and duration of EGFR-TKIs combination therapy (eg, 3 months, 3-6 months, 6-9 months). the interval between taking herbal medicines (e.g., continuous or intermittent), the discovery of different stages of NSCLC (e.g., 6 months <, 6 months >, 12 months >),

#### 2.7.2 Sensitivity analysis

Sensitivity analysis will be performed to determine the robustness and stability of the aggregated results by eliminating low-quality studies.

#### 2.7.3 Reporting bias analysis

If ≥ 10 randomized controlled trials will be included, a funnel chart will be performed to assess reporting bias.

### 2.8 Quality of evidence

We will use an improved Jadad scale to evaluate the quality of the literature in terms of random sequence generation, concealment of random allocation protocols, whether blinding is used in studies, case withdrawal and loss of follow-up, etc., with 1 to 3 being low quality and 4 to 7 being high quality.

### 2.9 Patient and Public Involvement

Patients or the public were not involved in the design, or conduct, or reporting, or dissemination plans of our research.

## 3. Ethics and dissemination

Because this systematic review will be conducted based on published research, there is no ethical approval requirement. The findings of this systematic review will be published in a peerreviewed journal.

## 4. Discussion

There is no relevant description of lung cancer in TCM, but according to the description of symptoms and signs, lung cancer can be classified as TCM cardiac, lung volume, lung wall and so on. Although there is no record of lung cancer disease names in the literature of TCM, the works of successive generations of doctors have accurate and detailed descriptions of the clinical symptoms of lung cancer, and some effective treatment methods have been established, which is still of guiding significance for the diagnosis and treatment of lung cancer in contemporary TCM. TCM is also an important treatment method for advanced lung cancer, and has good advantages in increasing efficiency and reducing toxicity and delaying tumor resistance. ^[16, 18]^ The whole process of Western medicine treatment and Traditional Chinese medicine is coordinated, learning from each other’s strengths, focusing on different stages, and timely adjusting the imbalance between yin and yang, which can not only alleviate various toxic side effects of Western medicine treatment, ensure the smooth progress of the treatment process, but also use TCM to enhance physical fitness. Patients with better physical condition are more likely to benefit from chemotherapy,^[19]^ so TCM can assist Western medicine in taking advantage of the chase and better prevent recurrence and metastasis of lung cancer. Therefore, the purpose of this systematic review is to determine the safety and efficacy of TCM in the treatment of NSCLC positive for EGFR mutations in combination with EGFR-TKIs, and to provide reference ideas for delaying the treatment of secondary drug resistance and drug resistance. Provide convincing evidence of efficacy to clinical practitioners, scientific researchers, and even patients in general.

## Data Availability

Data sharing not applicable to this article as no datasets were generated or analyzed during the current study.

## 5. Author contributions

Conceptualization: Shanyang Su, Yanling Huang,Wenjia Li, Xiang Lu,Xiaowan Chen,Jihong Zhou.

Data curation: Wenjia Li, Jihong Zhou.

Formal analysis: Shanyang Su,Yanling Huang.

Methodology: Shanyang Su,Yanling Huang,Jihong Zhou..

Project administration:. Xiang Lu,Xiaowan Chen,Jihong Zhou.

Resources: Shanyang Su, Jihong Zhou.

Software: Shanyang Su, Yanling Huang.

Visualization: Xiang Lu,Wenjia Li.

Writing – original draft: Shanyang Su,Yanling Huang,Wenjia LiWriting – review & editing: Xiaowan Chen, Jihong Zhou, Wenjia Li

